# Time-Varying COVID-19 Reproduction Number in the United States

**DOI:** 10.1101/2020.04.10.20060863

**Authors:** Douglas D. Gunzler, Ashwini R. Sehgal

## Abstract

The basic reproduction number is the average number of people to whom an infected person transmits the infection when virtually all individuals in a population are susceptible. We sought to calculate the current reproduction number for COVID-19 for each state in the United States. For the entire United States, the time-varying reproduction number declined from 4.02 to 1.51 between March 17 and April 1, 2020. We also found that the time-varying reproduction number for COVID-19 has declined in most states over the same two week period which suggests that social isolation measures may be having a beneficial effect.

## Introduction

The basic reproduction number is the average number of people to whom an infected person transmits the infection when virtually all individuals in a population are susceptible. A reproduction number greater than 1.0 means an epidemic is growing while a number less than 1.0 means an epidemic is slowing down and may eventually end. The reproduction number for the novel coronavirus responsible for COVID-19 was estimated to be 2.2 in Hubei Province, China, based on cases in December 2019 and January 2020.(1) Following one month of social distancing and lockdown, the reproduction number decreased to 1.6.(2) We sought to calculate the current reproduction number for each state in the United States.

## Methods

We used data on the number of daily cases as compiled by the New York Times from federal, state, and local sources to estimate reproduction number for the most recent day, one week earlier, and two weeks earlier.(3) This estimation requires information on the distribution of the serial interval, i.e. the time between symptom onset in an infector-infectee pair. We utilized serial interval data reported in China and truncated negative values and values greater than 20 days as unrealistic.(4) We identified the best function as a gamma distribution with a mean of 4.5 and a standard deviation of 4.3 days and calculated time dependent daily reproduction numbers over a 14 day interval using the approach proposed by Wallinga & Teunis with a Bayesian modification for real-time estimation.(4-6) All analyses were performed with R statistical software and R0 and projections packages.

## Results

For the entire United States, the reproduction number declined from 4.02 to 1.51 over the last two weeks (Table). Individual states varied greatly in their current reproduction number and trajectory over the last two weeks. For example, Ohio’s reproduction number declined from 3.97 to 1.62. By contrast, Louisiana had a more gradual decline and had a reproduction number on April 1 of 2.22.

**Table.**
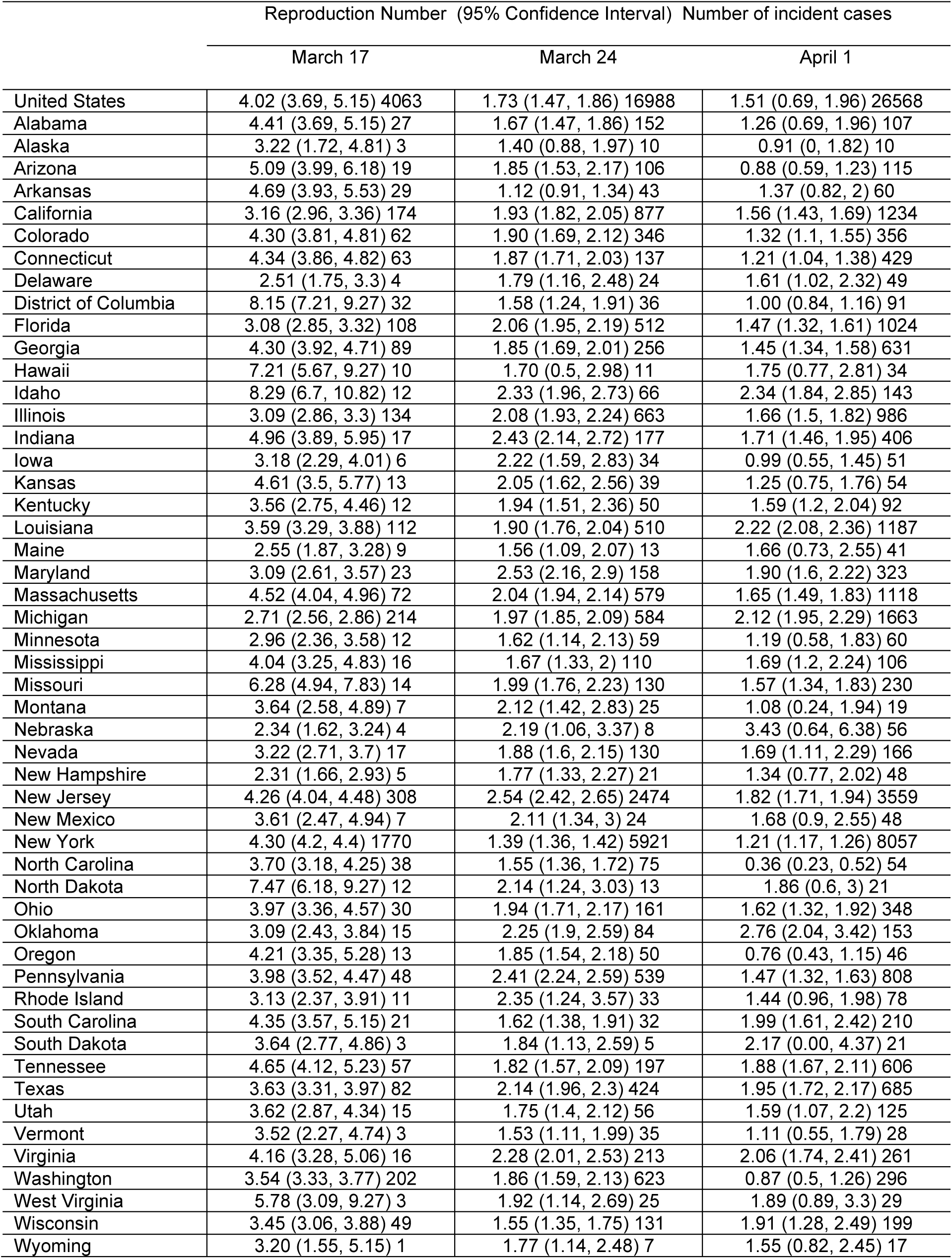
Time varying COVID-19 reproduction number over the last two weeks in the United States.

Even small differences in reproduction number can have sizeable effects. For example, Ohio will be projected to have 3500 incident cases in three weeks if the current reproduction number of 1.62 persists. The projected number of cases would be 1400 with a reproduction number of 1.40 and 6500 with a reproduction number of 1.80.

## Discussion

Several factors influence the value of the reproduction number, including how contagious an infectious organism is, how many susceptible people there are and their proximity to each other, and what measures are implemented to limit spread. We found that the reproduction number for COVID-19 has declined in most states over the past two weeks which suggests that social isolation measures may be having a beneficial effect. However, most states still have reproduction numbers substantially higher than 1.0. That means it would be premature to relax restrictions and resume normal social and economic activities. Doing so now will result in an increase in the reproduction number and a surge of new cases. Examining changes in reproduction number over the coming weeks and months may help guide decisions to continue social restrictions and to monitor the impact of gradually relaxing such restrictions.

Many COVID-19 cases, particularly asymptomatic and mild ones, are not reported to public health departments so our calculation may underestimate the true reproduction number. In addition, the number of people tested has increased greatly in recent days as testing kits have become more available. This would be expected to increase COVID-19 diagnoses and therefore the reproduction number. It is encouraging that we found a reduction in reproduction number despite this increase in testing. Other limitations include the lack of United States data on serial intervals, daily reproduction numbers can be highly variable and the small numbers of cases in some states.

## Data Availability

We used data on the number of daily cases as compiled by the New York Times from federal, state, and local sources to estimate reproduction number for the most recent day, one week earlier, and two weeks earlier.
New York Times COVID-19 data. Available at https://github.com/nytimes/covid-19-data Accessed April 2, 2020.

## Acknowledgments

This study was supported in part by grants UL1 TR002548 and U54 MD002265 from the National Institutes of Health.

